# The relationship between blastocyst morphology and the sex ratio of singleton births in single blastocyst transfer cycles: A retrospective cohort study

**DOI:** 10.1101/2022.11.28.22282845

**Authors:** Jijun Hu, Yichun Guan, Rujing Yang, Lijun Sun, Hua Lou, Xingling Wang

## Abstract

**Background:** To study the relationship between blastocyst morphology and the sex ratio of singleton births in single blastocyst transfer cylces.

**Methods:** This retrospective cohort study included women who underwent *in vitro* fertilization/intracytoplasmic sperm injection-embryo transfer (IVF/ICSI-ET) from January 2017 to December 2019 and obtained a single live baby following a single blastocyst transfer. The sex ratio which influenced by many possible factors were studied. Age-stratified analysis was performed for the maternal and paternal groups younger and older than 35 years. The 3108 cycles included here were grouped by sex at birth (male: 1777 cycles and female: 1331 cycles). Blastocysts were conceived via 2834 IVF and 274 ICSI treatments, and were used for 1054 fresh ET cycles and 2054 frozen-thawed embryo transfer (FET) cycles.

**Results:** Sex ratio among singleton of poor blastocyst decreased significantly compared to those of good blastocyst (51.5% vs 60%, *P*<0.001). The results of multivariate logistic analysis showed that high-quality blastocysts had a 151% higher probability of being male singleton than poor-quality blastocysts (*P*<0.001). Further stratified analysis shows that in both group younger than 35 the sex ratio increased significantly in grades A and/or B trophectoderm (*P*<0.001) and good-quality blastocysts (*P*<0.001). While grade A inner cell mass (ICM) degree was significantly associated with a lower sex ratio than grade B (*P*<0.05).

**Conclusions:** A single high-quality blastocyst transfer was significantly associated with a higher neonatal sex ratio. Grade A trophectoderm and grade B ICM contributed to improving the implantation potential of male blastocysts, especially in infertile men or women ≤35 years old.

## Background

IVF was first developed over 40 years ago. Millions of babies have since been born as a result of infertility treatment worldwide. Routine blastocyst culture and transfer using sequential culture media in IVF/ICSI is associated with a significant increase in implantation rates and clinical pregnancy rate.However,there are some adverse effects in blastocyst transfer such as a male-biased imbalance in the sex ratio of newborns and an increased incidence of monozygotic twinning (MZT). [1-3]

A three-part scoring system,which based on blastocyst expansion, inner cell mass, and trophectoderm development formulated by Gardner and Schoolcraft [4], aiming to grade human blastocysts before transfer. The scoring system brought great convenience to embryologists and researchers’ studies on the development and viability of blastocyst.In many cases, blastocysts achieving the best morphologic scores often achieve implantation and produce a live birth,and those with high scores on trophectoderm have relatively easy access to a male child (2021) .

The human sex ratio is often expressed as the number of males per 100 female live births [5]. In Western gender-neutral countries, the sex ratio at birth is estimated to be approximately 1.06 [6]. In Japan, the overall neonatal sex ratio has steadily declined from 1.079 in 1970 to as low as 1.056 in 2016 (Ministry of Health, Labour and Welfare Vital Statistics of Japan 2016). In the 2000 census in China, the reported sex ratio at birth was 117 per 100 female births, which appears to be due to a declining fertility rate and a traditional preference for males, a phenomenon that is common to many other Asian countries [7]. A recent study by Fan et al. reported that the sex ratio at birth gradually declined to normal levels (1.047 with the universal child policy) following the gradual rollout of the two-child policy in China [8].

The sex ratio at birth from assisted artificial technology cycles ranges from 1.03–1.25. However, there are discrepancies in the sex ratio between embryos generated by standard IVF/ICSI, and between cleavage and single blastocyst transfer (SBT) embryos. As SBT is the most effective approach for avoiding multiple pregnancies, SBT has gradually become the preferred embryo transfer strategy throughout the world [9-14]. However, a male-biased imbalance in the sex ratio is one of the important potential limitations in blastocyst transfer [1, 3]. Several studies have reported a significantly higher sex ratio at birth among infants born after SBT (54.1%) than among those born after cleavage-stage (day 3) transfer cycles (49.9%) [15, 16].

In practice, embryologists tend to select preferentially better morphology and more developmentally advanced blastocysts for transfer, because good-quality blastocyst has higher implantation rate and clinical pregnancy rate. The contribution of these parameters to the implantation potential or sex ratio is not fully known. Some studies have reported that trophectoderm morphology may be predictive of pregnancy outcomes and sex ratio related to SBT [17, 18].

We intended to study the relationship between blastocyst morphology and the sex ratio of singleton births in single blastocyst transfer. To that end, here we analyzed clinical data for the SR of IVF/ICSI children conceived after SBT between 2017 and 2019. The variables considered in this research included: assisted reproductive technologies (ART) such as IVF, ICSI, FET, or ET, maternal or paternal age, and year of infertility.

## Methods

### Study subjects

This retrospective cohort study included the singleton offspring born to mothers after successful treatment cycles following the autologous SBT of non-preimplantation genetic testing (PGT) blastocysts at the Reproductive Medicine Center of the Third Affiliated Hospital of Zhengzhou University between January 2017 and December 2019. The study was carried out after the protocol was approved by the Institutional Review Board of The Third Affiliated Hospital of Zhengzhou University. Informed consent was waived due to retrospective design of the study based on our local ethics committee protocols.

### Ovarian stimulation protocol

Each female patient underwent a conventional ovarian hyperstimulation procedure [19, 20] involving a gonadotrophin releasing hormone agonist or antagonist or mild stimulation protocol. The physician adjusted the starting dose according to the patient’s age, body mass index (BMI) and ovarian reserve. Ovarian follicle development was monitored based on serum estradiol and transvaginal ultrasonographic measurements. Oocytes were retrieved transvaginally 36–38h after human chorionic gonadotrophin (Serono, Aubonne, Switzerland) administration when at least 40% of the follicles had reached or exceeded an average diameter of 18mm as determined by ultrasound. The follicles were aspirated using a single-lumen needle attached to a syringe under transvaginal ultrasound guidance. The oocytes were then inseminated via conventional IVF/ICSI.

### Laboratory protocol

Embryos were placed in an incubator (K-MINC-1000, Cook, USA) and cultured at 37°C under 6% CO_2_ and 5% O_2_. Embryos were cultured in G-1™ plus (Vitrolife, Sweden) from the pronucleate stage to day 3. On day 3 of culture, the embryo cell number and morphology were scored. Day 3 embryos are scored from I to IV. Classes I–III represent embryos that are available for transfer. After scoring, the embryos were subsequently transferred to G-2™ plus (Vitrolife, Sweden) medium for an additional 2–3 days of culture (days 5–6 of development) [21]. On day 5, the percentage of blastocyst formation was determined, and one blastocyst was selected for fresh cycle transfer, and the untransferred blastocysts were cryopreserved. On day 5 or 6 after insemination, blastocyst morphology was evaluated according to that described by Gardner and Schoolcraft [22]. Blastocysts expansion degree > 3, inner cell mass (ICM) grade A/B, and trophectoderm grade A/B (AA, AB, BA, BB) were considered high-quality blastocysts. Blastocysts expansion degree > 3, ICM grade A/B and trophectoderm grade C (AC, BC) were considered low-quality blastocysts [23]. All gamete and embryo manipulations occurred in a clean bench designed to control humidity, temperature, and pH fluctuation.

A laser hole caused the blastocysts to collapse, and the shrunken blastocyst was vitrified using a Cryotop device (Kitazato BioPharmao. Shizuoka, Japan). Vitrification and warming kits were used as media. For vitrification, the blastocysts were first equilibrated in solution I (7.5% v/v ethylene glycol [EG] and 7.5% v/v dimethyl sulfoxide [DMSO]) at room temperature for 10 min and then placed into vitrification solution II (15% v/v EG, 15% v/v DMSO, and 0.5 M sucrose) for 1 min. Subsequently, blastocysts were individually loaded onto Cryotop in a volume of <0.1μL and quickly plunged into liquid nitrogen. The blastocyst was warmed by placing the Cryotop in thawing solution (1.0 M sucrose) for 1 min at 37°C. The blastocyst was then transferred to dilution solution (0.5 M sucrose) for 3 min at room temperature, then washed twice in washing solution at room temperature for 3 min each. The blastocyte was transferred into a 20-μL droplet of culture medium (G-2; Vitrolife) under mineral oil. Blastocysts were transferred 1–2 h post-warming.

### Embryo transfer

Fresh single blastocyst transfer was performed 5 days after fertilization in our Reproductive Center. ET was routinely performed under abdominal ultrasound guidance. Fresh ET was cancelled in cases with severe ovarian hyperstimulation syndrome (OHSS), an endometrial thickness of ≤7 mm, progesterone levels ≥2μg/L on the human chorionic gonadotropin (HCG) trigger day, and some other special conditions; all the embryos were cryopreserved and FETs were subsequently performed to decrease the rate of OHSS and to improve the clinical pregnancy rate.

FET and clinical pregnancy confirmed were performed referring to the descriptions of the previous study[17].The singleton SR was primary outcome measure of this study. The sex ratio at birth was calculated as the number of males per 100 female live births.

### Statistical analysis

All data analyses were performed using Statistical Package for Social Sciences (SPSS) software package version 22.0 (IBM Corp, Armonk, NY, USA). For continuous variables, normally distributed variables are expressed as the mean and standard deviation, while non‐normally distributed variables are shown as the median and interquartile range. Categorical variables are summarized as frequencies with proportions. The unpaired Student’s *t* test or Mann–Whitney *U* test was used for comparing the difference between the two groups. Univariate regression analysis was performed for factors, including maternal age, paternal age, mean duration of infertility, mean maternal BMI, type of infertility, type of infertility (%), day of blastocyst transfer, type of fertilization, quality of transferred blastocyst, ICM, and trophectoderm grade. Both crude and adjusted multivariable logistic regression models for sex ratio of live singleton were established (enter). For the adjusted multivariate logistic regression model, factors including maternal age, paternal age, BMI, duration of infertility, type of infertility, and day of blastocyst transfer were further adjusted. *P* value <0.05 was set as the threshold of statistical significance.

## Results

### Study participants and baseline characteristics

A total of 5632 SBT cycles of women who underwent IVF/ICSI-ET were screened. Of these, 177 cycles that underwent PGT were excluded. Of the 5285 autologous cycles remaining, there were 328 miscarriages, 29 ectopic pregnancies, and 56 cycles that resulted in monozygotic twins. After excluding these cycles, there were 3108 cycles that resulted in singleton live births, which were included in the analysis. There were 1054 (33.9%) fresh blastocyst transfer cycles and 2054 (66.1%) frozen blastocyst transfer cycles,2070 (66.6%) high-quality and 1038 (33.4%) poor-quality single blastocyst FETs. The blastocysts were conceived via 2834 (91.2%) IVF and 274 (8.8%) ICSI treatments. The analyzed cycles yielded 1777 male infants and 1331 female infants, and the sex ratio was 133% (133/100). All the SBT cycles of each patient was analyzed to avoid repeated measures bias. Patients who underwent PGT or used donor oocytes or donor sperm were excluded from this study. Figure 1 presents the identification and collation process of the study sample. Table 1 describes the baseline characteristics of the included treatment cycles.

**Table 1.** Characteristics of 3108 cycles, including sex ratio, among singleton births from January 2017 to December 2019.

| Live birth sex | Female | Male | P value |
| --- | --- | --- | --- |
| N | 1331 | 1777 |  |
| Maternal age, years (mean ± SD) | 30.29 ± 4.32 | 30.12 ± 4.25 | 0.288 |
| Paternal age, years (mean ± SD) | 31.30 ± 5.05 | 31.19 ± 5.02 | 0.578 |
| Mean duration of infertility (years ± SD) | 3.11 ± 2.48 | 3.17 ± 2.50 | 0.511 |
| Mean maternal BMI (kg/m<sup>2</sup>) | 23.80 ± 3.27 | 23.58 ± 3.21 | 0.061 |
| Type of infertility (%) |  |  | 0.605 |
| Primary | 604 (45.38%) | 823 (46.31%) |  |
| Secondary | 727 (54.62%) | 954 (53.69%) |  |
| Factor of infertility (%) |  |  | 0.881 |
| Female factor | 687 (51.62%) | 929 (52.28%) |  |
| Male factor | 324 (24.34%) | 436 (24.54%) |  |
| Mixed factor | 240 (18.03%) | 301 (16.94%) |  |
| Unexplained infertility | 80 (6.01%) | 111 (6.25%) |  |
| Type of blastocyst transfer (%) |  |  | 0.073 |
| ET | 428 (32.16%) | 626 (35.23%) |  |
| FET | 903 (67.84%) | 1151 (64.77%) |  |
| Day of blastocyst transfer (%) |  |  | 0.655 |
| D5 | 1039 (78.06%) | 1399 (78.73%) |  |
| D6 | 292 (21.94%) | 378 (21.27%) |  |
| Type of fertilization (%) |  |  | 0.832 |
| ICSI | 1212 (91.06%) | 1622 (91.28%) |  |
| IVF | 119 (8.94%) | 155 (8.72%) |  |
| Blastocyst quality of transfer (%) |  |  | **<0.001** |
| Poor | 503 (37.79%) | 535 (30.11%) |  |
| Good | 828 (62.21%) | 1242 (69.89%) |  |
| ICM |  |  | **0.007** |
| A | 414 (31.10%) | 474 (26.67%) |  |
| B | 917 (68.90%) | 1303 (73.33%) |  |
| TE |  |  | **<0.001** |
| A | 261 (19.61%) | 418 (23.52%) |  |
| B | 567 (42.60%) | 824 (46.37%) |  |
| C | 503 (37.79%) | 535 (30.11%) |  |

Note: The sex ratio at birth is calculated as the proportion of males among all livebirths.
CI, confidence interval; BMI, body mass index; D, day; IVF, in vitro fertilization; ICSI, intracytoplasmic sperm injection; ET, fresh embryo transfer; FET, frozen embryo transfer

**Figure. 1.**
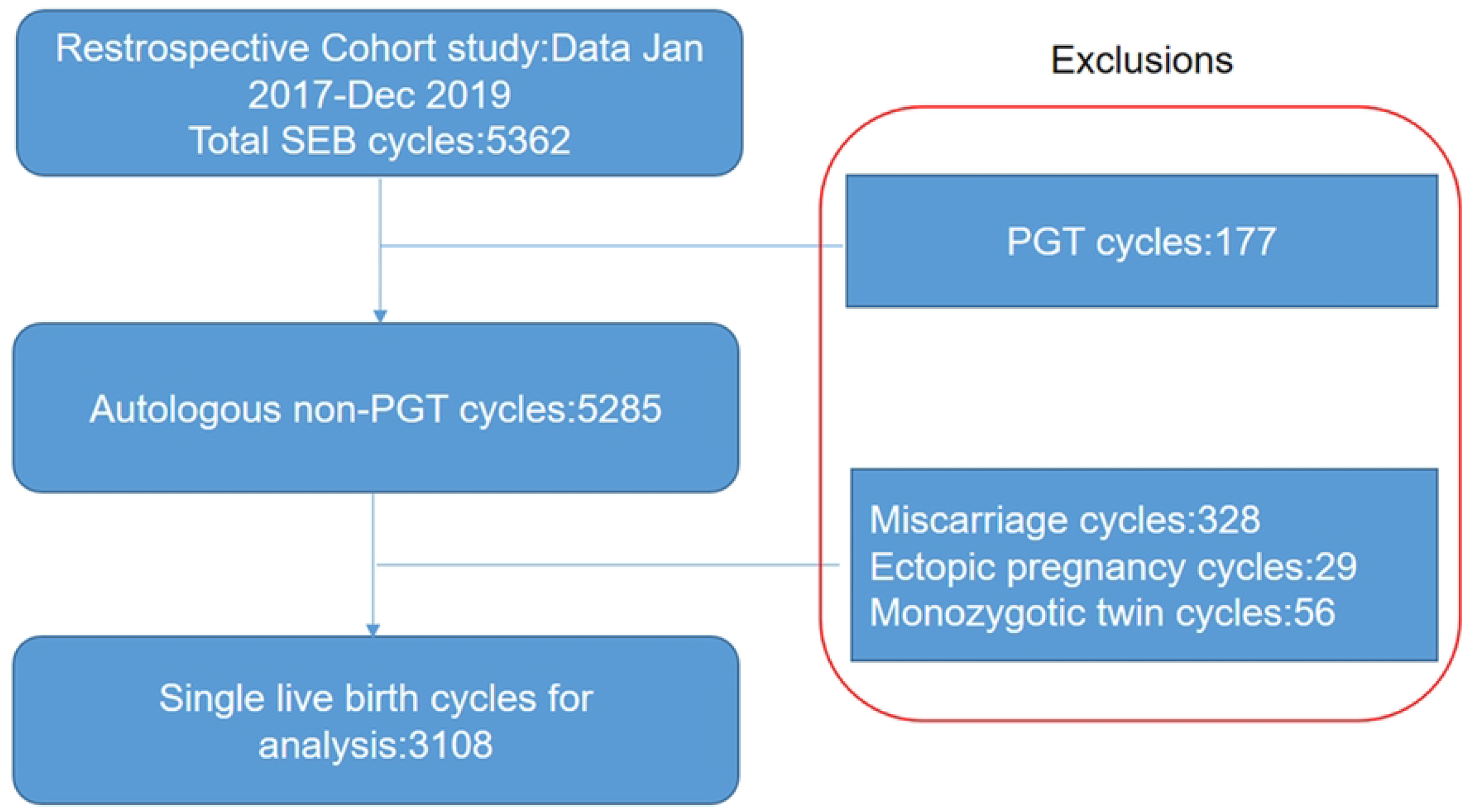
Data selection process for the analysis of perinatal outcomes in singleton live births resulting from single blastocyst transfer (SBT) cycles Note: Flowchart showing exclusions made to singleton cycles for analysis data to identify component subsets that were merged to construct the final study dataset. PGT=preimplantation genetic testing

The results of the univariable analysis for the potential factors influencing sex ratio among singleton live births revealed that transfer of a single high-quality blastocyst resulted in a significantly higher sex ratio than SBT of a poor-quality blastocyst (60% vs. 51.5%, *P* < 0.001, Fig. 2). Transfer of blastocysts with a higher TE grade and lower ICM grade resulted in a significantly higher sex ratio. Maternal age, paternal age, duration of infertility, maternal BMI, main infertility cause, type of fertilization, or day of embryo transfer had no significant influence on the sex ratio (Table 2).

**Table 2.**
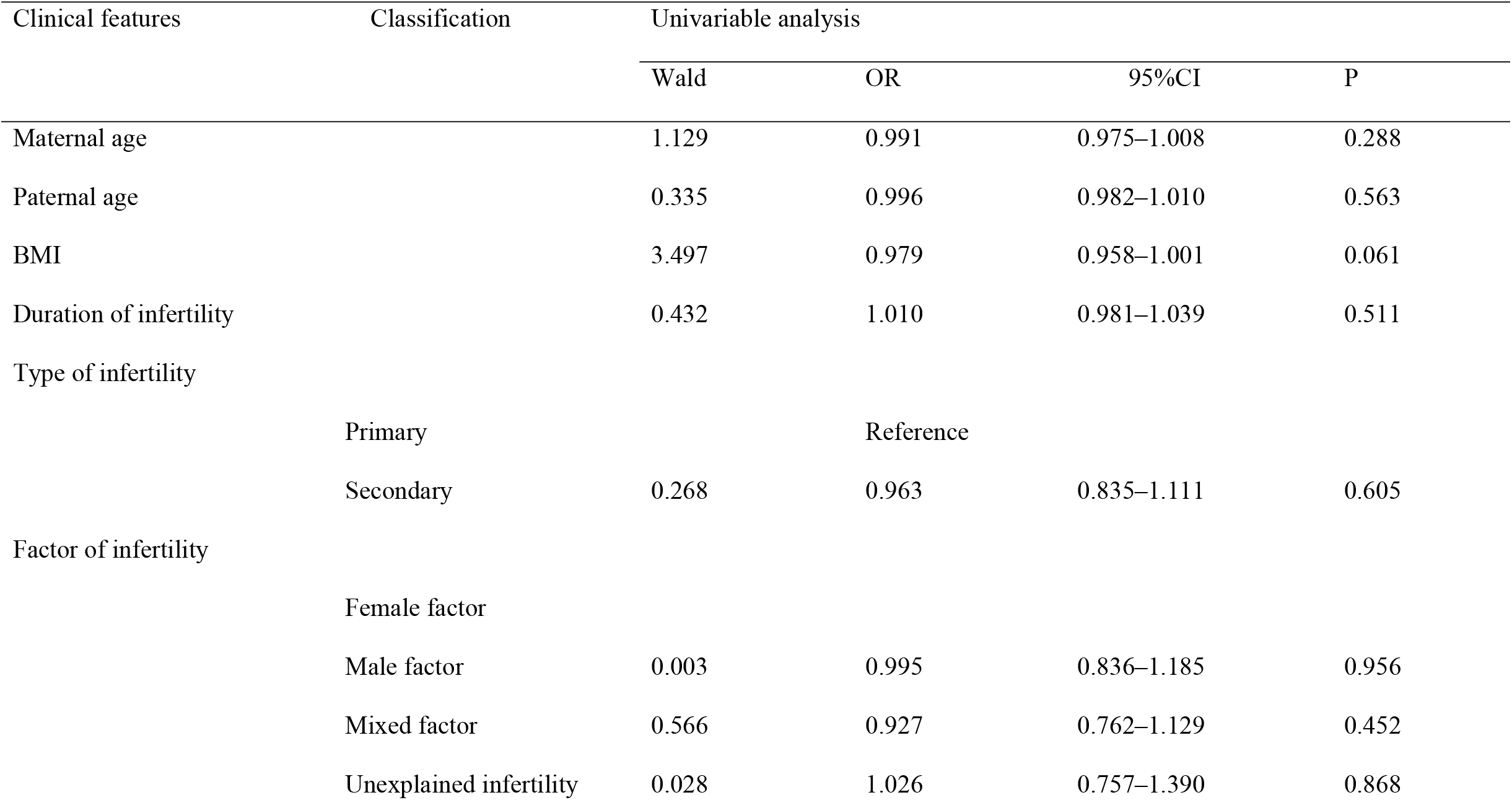
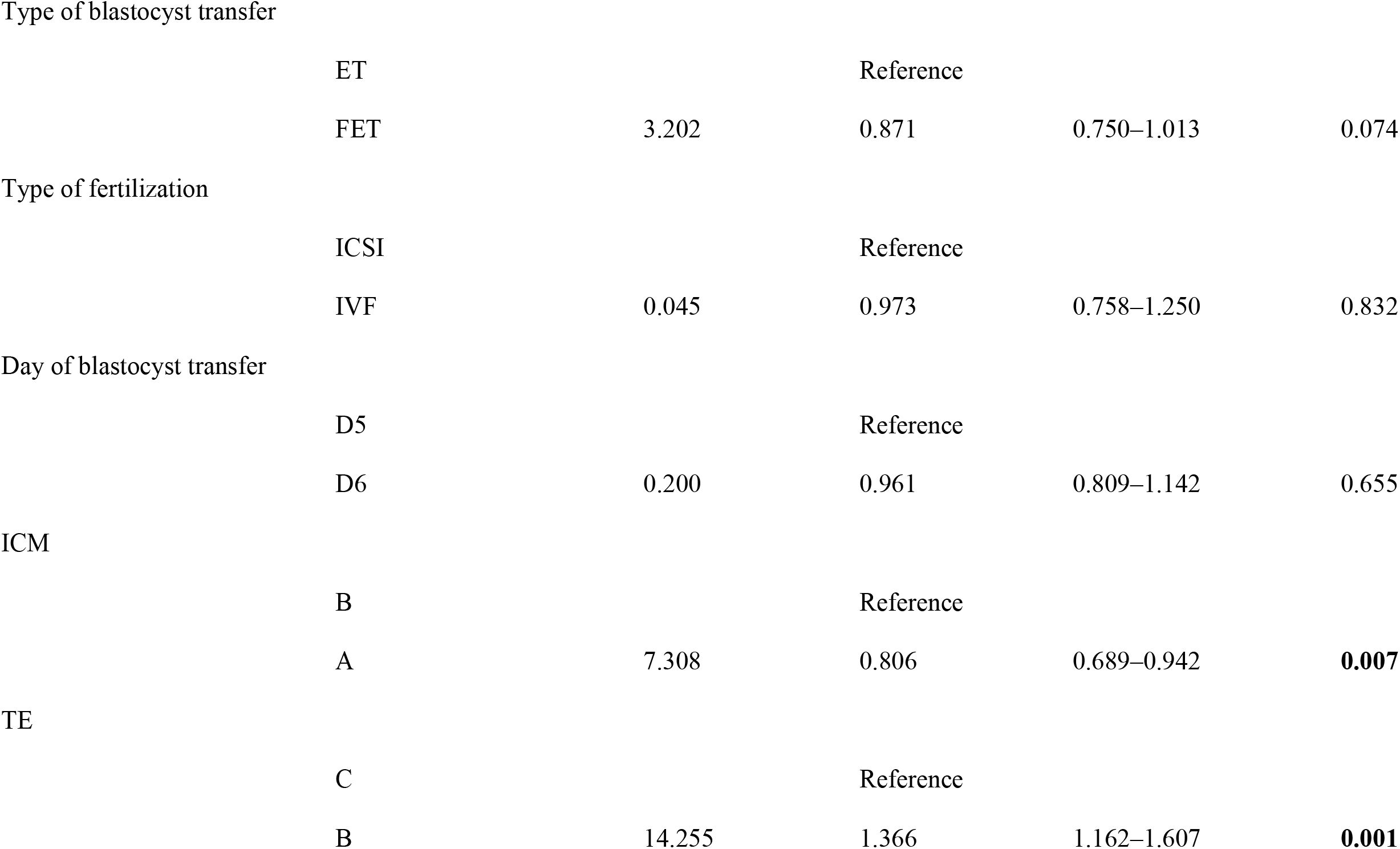
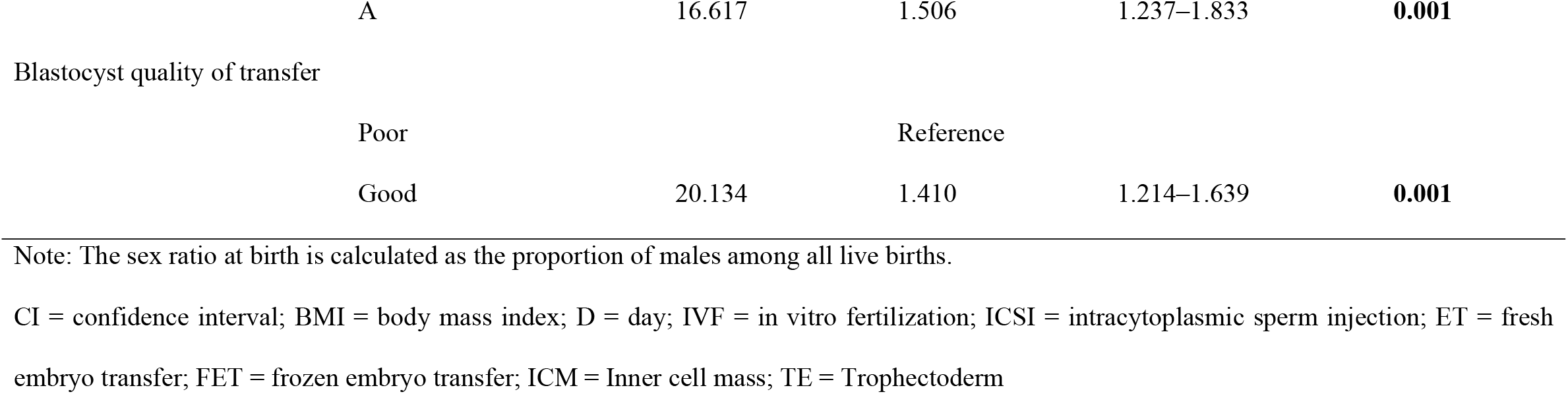
Univariable analysis for the potential factors influencing sex ratio among singleton live births

| Clinical features | Classification | Univariable analysis |  |  |  |
| --- | --- | --- | --- | --- | --- |
|  |  | Wald | OR | 95%CI | P |
| Maternal age |  | 1.129 | 0.991 | 0.975–1.008 | 0.288 |
| Paternal age |  | 0.335 | 0.996 | 0.982–1.010 | 0.563 |
| BMI |  | 3.497 | 0.979 | 0.958–1.001 | 0.061 |
| Duration of infertility |  | 0.432 | 1.010 | 0.981–1.039 | 0.511 |
| Type of infertility |  |  |  |  |  |
|  | Primary |  | Reference |  |  |
|  | Secondary | 0.268 | 0.963 | 0.835–1.111 | 0.605 |
| Factor of infertility |  |  |  |  |  |
|  | Female factor |  |  |  |  |
|  | Male factor | 0.003 | 0.995 | 0.836–1.185 | 0.956 |
|  | Mixed factor | 0.566 | 0.927 | 0.762–1.129 | 0.452 |
|  | Unexplained infertility | 0.028 | 1.026 | 0.757–1.390 | 0.868 |
| Type of blastocyst transfer | ET |  | Reference |  |  |
|  | FET | 3.202 | 0.871 | 0.750–1.013 | 0.074 |
| Type of fertilization | ICSI |  | Reference |  |  |
|  | IVF | 0.045 | 0.973 | 0.758–1.250 | 0.832 |
| Day of blastocyst transfer | D5 |  | Reference |  |  |
|  | D6 | 0.200 | 0.961 | 0.809–1.142 | 0.655 |
| ICM | B |  | Reference |  |  |
|  | A | 7.308 | 0.806 | 0.689–0.942 | **0.007** |
| TE | C |  | Reference |  |  |
|  | B | 14.255 | 1.366 | 1.162–1.607 | **0.001** |

|  |  |  |  |  |
| --- | --- | --- | --- | --- |
| A | 16.617 | 1.506 | 1.237–1.833 | **0.001** |
| Blastocyst quality of transfer |  |  |  |  |
| Poor |  | Reference |  |  |
| Good | 20.134 | 1.410 | 1.214–1.639 | **0.001** |

Note: The sex ratio at birth is calculated as the proportion of males among all live births.
CI = confidence interval; BMI = body mass index; D = day; IVF = in vitro fertilization; ICSI = intracytoplasmic sperm injection; ET = fresh embryo transfer; FET = frozen embryo transfer; ICM = Inner cell mass; TE = Trophectoderm

**Figure. 2.**
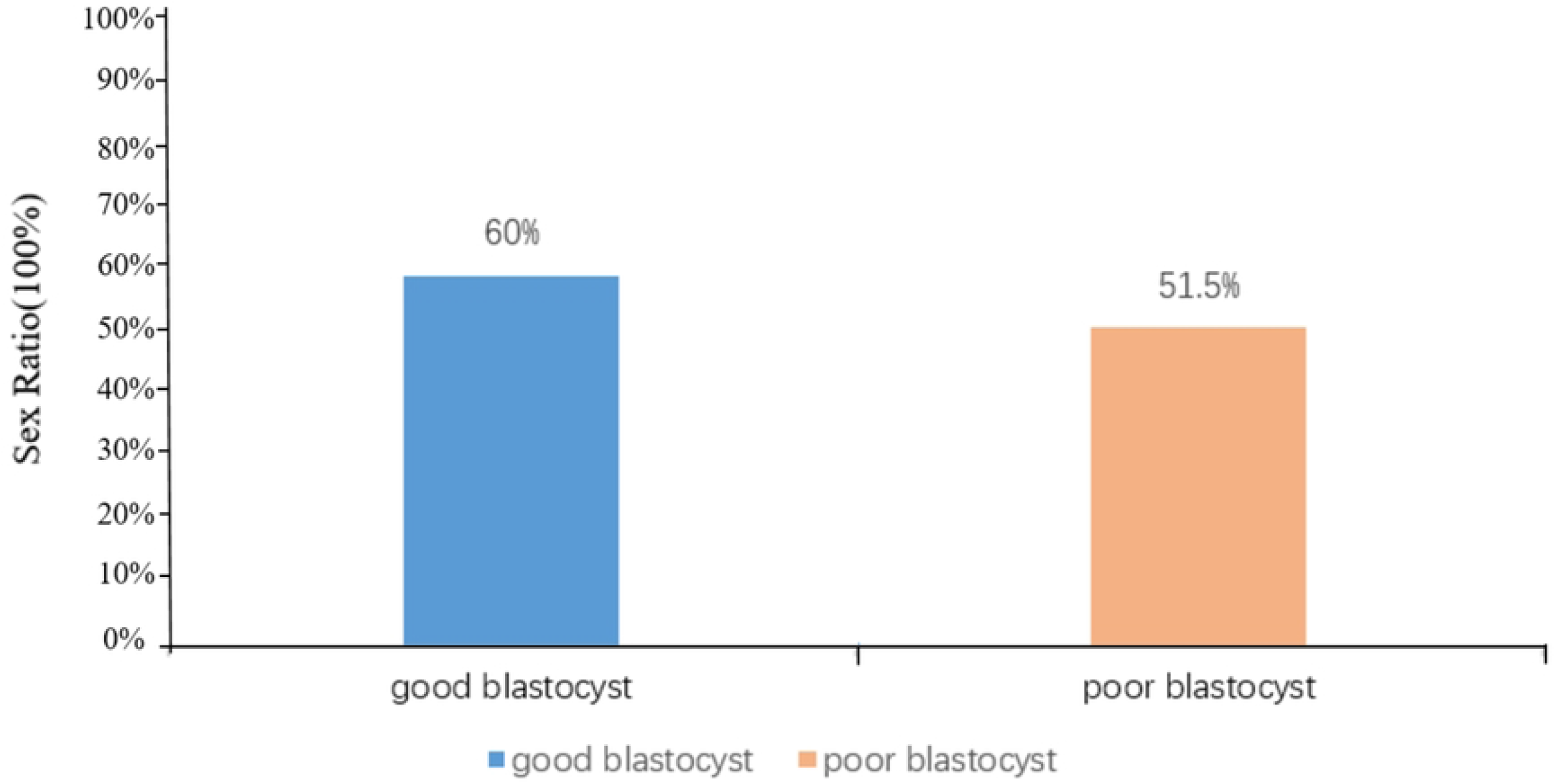
Sex ratio among singleton births following single blastocyst transfer (SBT) cycles with different blastocyst qualities. Note: The sex ratio at birth is calculated as the proportion of liveborn males per 100 females. CI = confidence interval; P value* was the P value after adjustment (P<0.001).

### Multivariate analysis

Multivariate analysis was performed after adjusting for potential confounding factors. Based on the results of the analysis, we identified significant associations of sex ratio among singleton births with the morphological characteristics of blastocysts.

The results of the multivariate analysis revealed that high-quality blastocysts have a 190% higher probability of being male than poor-quality blastocysts (aOR, 1.964; 95% CI, 1.559–2.474; *P* < 0.001). However, blastocysts with grade A ICM had a 37% lower probability of being male than in blastocysts with grade B trophectoderm (aOR 0.632; 95% CI, 0.529–0.757; *P* < 0.001). Moreover, when the trophectoderm was of grade A, there was a 190% higher probability of being male than when the trophectoderm was of grade C (aOR, 1.964; 95% CI, 1.559–2.474; *P* < 0.001; Table 3).

**Table 3.**
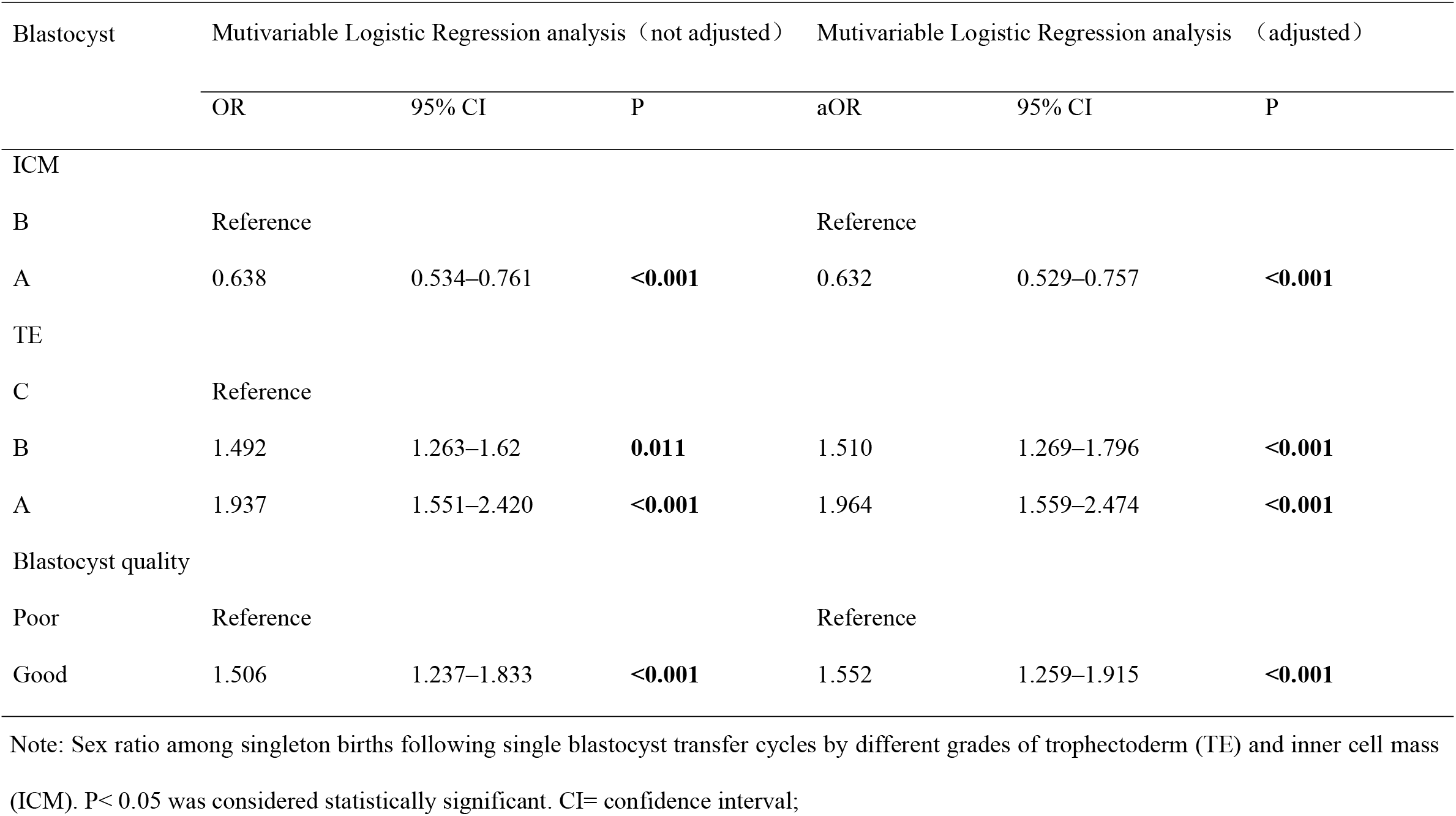
Multivariate logistic regression model of sex ratio (male) among singleton births following single blastocyst transfer cycles in relation to blastocyst quality and morphology.

### Stratified analysis by maternal and paternal age

We also determined relationship between quality classification of ICM grade, TE grade of transferred blastocysts and sex ratio according to parents aged 35 and older. When the maternal age was ≤35, blastocysts with grade A ICM had a significantly lower likelihood of being male than with grade B ICM (OR, 0.803; 95% CI, 0.677– 0.951; *P* = 0.011). Similarly, when the paternal age was ≤35 years, blastocysts with grade A ICM had a significantly lower likelihood of being male than blastocysts with grade B ICM (OR, 0.791; 95% CI, 0.663–0.942; *P* = 0.009). Blastocysts with grades A and B TE in women ≤35 years and grade B TE in men ≤35 years had a significantly higher sex ratio than those with grade C TE (women ≤35 years grade B TE: OR, 1.401; 95% CI, 1.172–1.676 and grade A TE: OR, 1.589; 95% CI, 1.282–1.970; *P* <0.001 for both) (men ≤35 years grade B TE: OR, 1.555; 95% CI, 1.245–1.942; *P* <0.001). Furthermore, good-quality blastocysts were significantly more likely to be male in both women and men aged ≤35 years (OR: 1.461, 95% CI, 1.238–1.725; *P* <0.001 for women and OR: 1.442; 95% CI, 1.214–1.712; and *P* <0.001 for men) (Table 4)

**Table 4.**
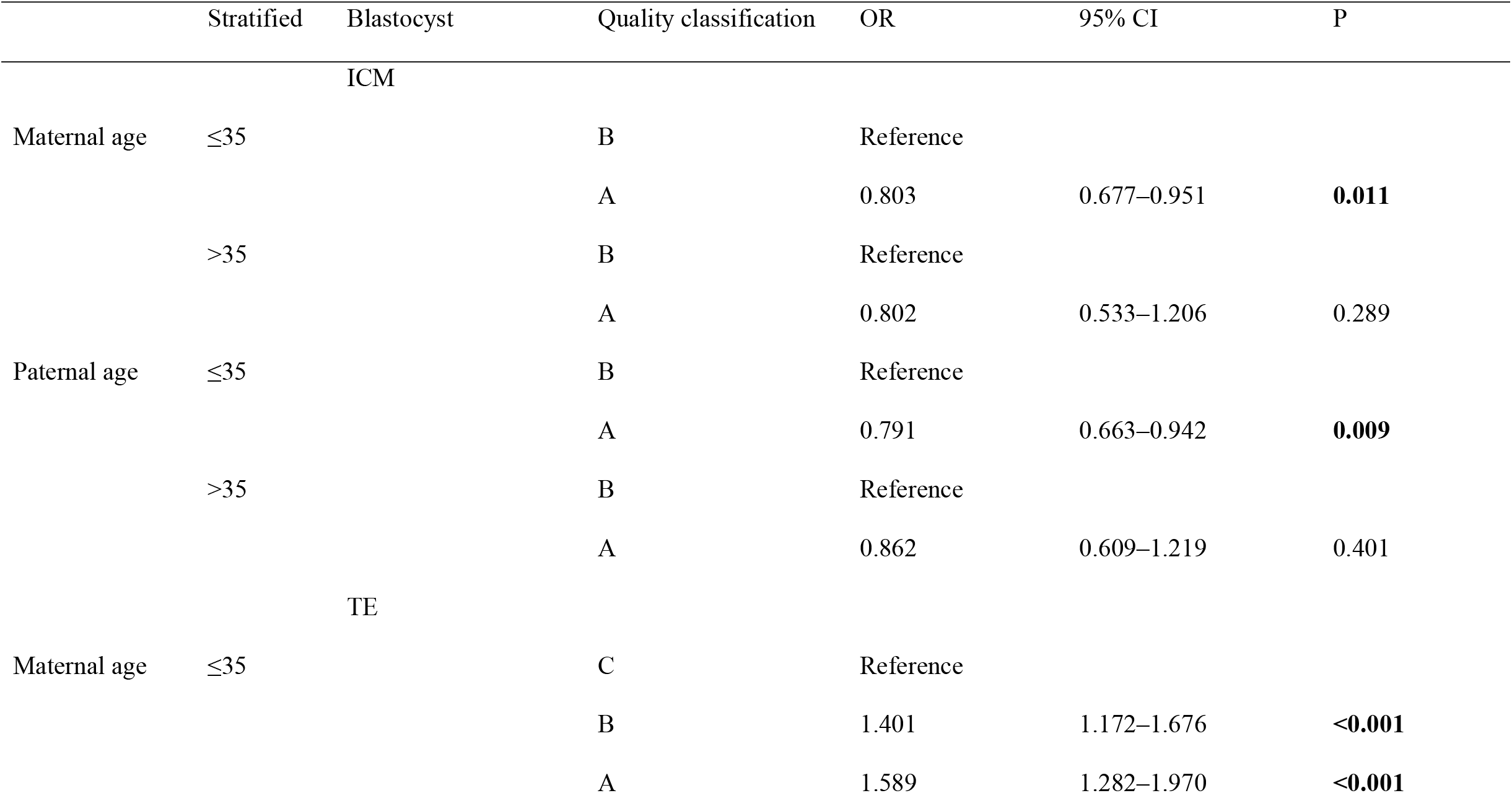
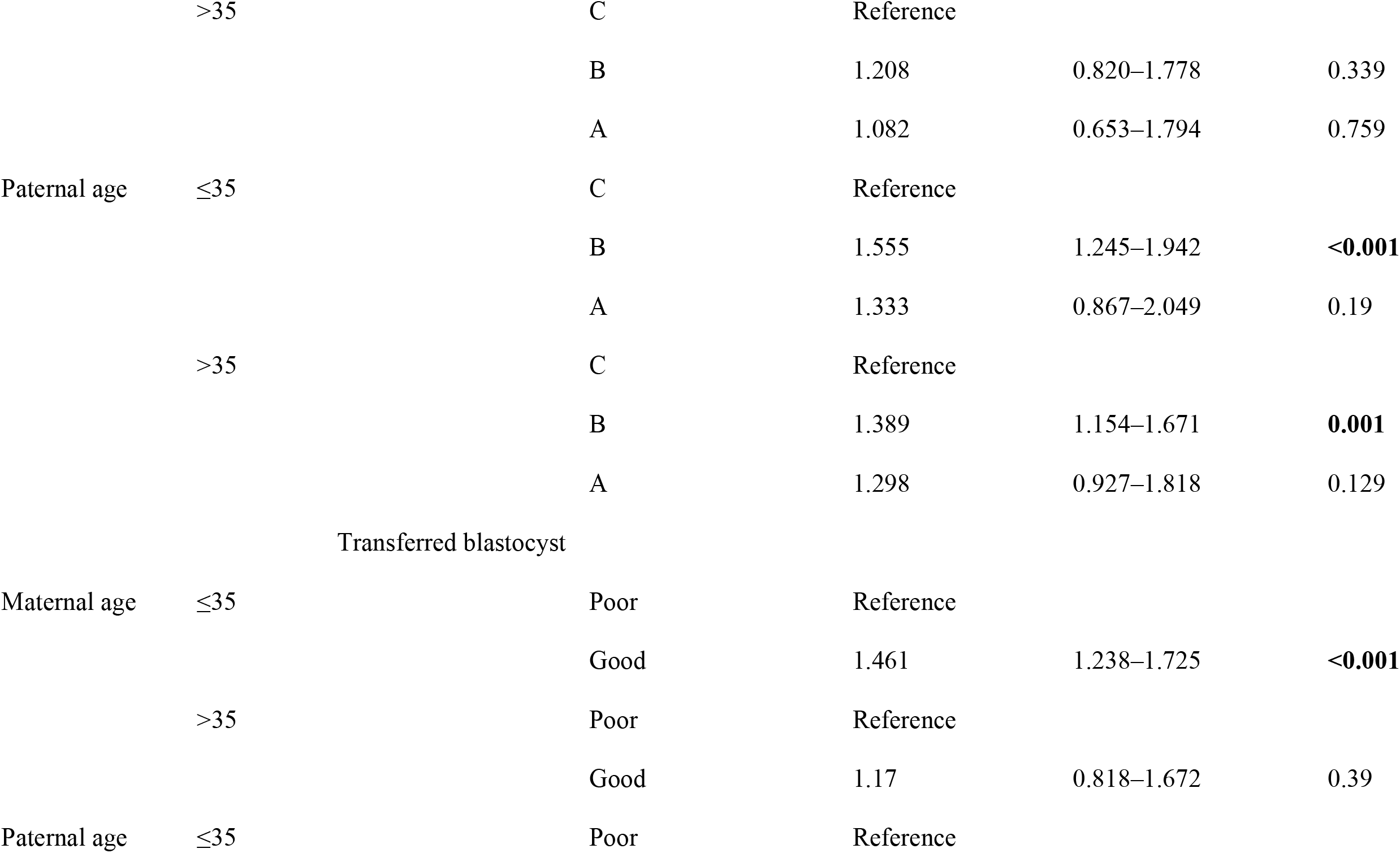
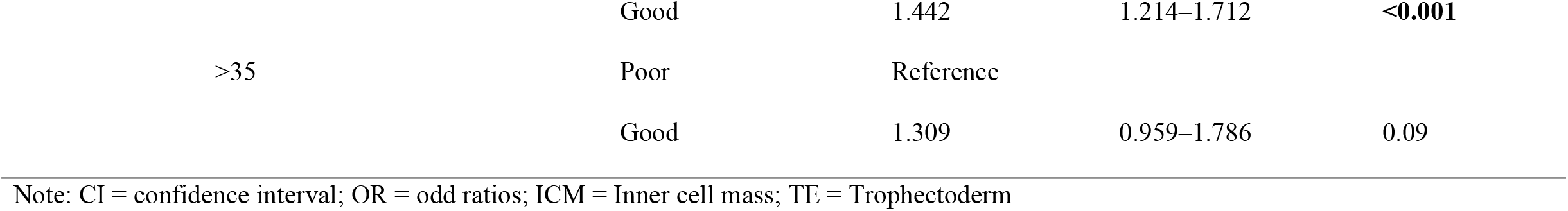
Effect of stratified maternal and paternal age and quality classification of ICM grade, TE grade, transferred blastocysts on sex ratio.

## Discussion

In the present study, SBT was used to avoid the complications associated with multiple pregnancies and maintain higher rates of implantation, pregnancy, and live birth in couples undergoing ART. Our results following the analysis of 3108 cycles revealed that high-quality blastocysts had a significantly higher probability of being male than poor-quality blastocysts. However, blastocyst culture conditions contributed to preferential selection of male blastocysts for transfer because of the advanced developmental rates *in vitro*. It is worthwhile to note that some studies explored the relationship between the stage at which ET occurs and the SSR, and found that ET at the blastocyst stage tends to increase the SSR compared with ET at the cleavage stage, and this effect is independent of other factors [24].

Among the 3108 IVF/ICSI cycles over a three-year period, there were 1777 males and 1331 females, making the SR 133 males per 100 females, which was consistent with the ratios reported in previous studies. A large nationwide cohort study in Japan showed a higher sex ratio after blastocyst transfer relative to spontaneous conception (OR: 1.095; 95% CI: 1.001–1.198) [3]. The study also found that implantation with high-quality blastocysts was associated with an increased sex ratio during SBT cycles. The similar result that blastocyst transfer was associated with increased sex ratio was also found in a retrospective cohort study [25].

We also explored the effect of FET on SSR. Very few studies have reported differences in the SSR following fresh and frozen ET, and the results are controversial [26, 27]. However, the results of our preliminary univariable analysis after controlling for related factors, including maternal age, paternal age, mean duration of infertility, mean maternal BMI, type of infertility, type of infertility (%), day of blastocyst transfer, type of fertilization, quality of transferred blastocyst, ICM, and trophectoderm grade, showed that the SSR was not related with FET (OR = 0.871, 95% CI: 0.750–1.013, *P* = 0.074). In most reproductive medical centers in China, blastocyst transfers are usually performed in FET cycles, and our center was no exception. It may be that blastocyst transfer and not FET is responsible for the skewing of the SSR toward males.

Previous studies have explored the effect of the IVF/ICSI insemination method on sex ratio. [28] ICSI, which is primarily used to treat male factor infertility, is now widely used all over the world. Data from other countries have shown that ICSI use significantly reduces the proportion of male babies compared with IVF (48.8% versus 51.4% in America; 50.0% vs 53.0% in Australia and New Zealand) [15, 29]. Although the sex ratio with ICSI insemination was found to be lower than that with IVF insemination in our study, this difference was not statistically significant. One study showed that ICSI reduces SSR only in cleavage-stage embryo transfer cycles, but not in blastocyst transfer cycles [24]. Currently, it is unclear why ICSI insemination reduces SSR. Tarín et al. compared sex ratios at implantation and birth in cleavage- and blastocyst-stage transfer cycles to propose molecular mechanisms accounting for differences in post-implantation mortality in male and female offspring and consequent variations in the sex ratios at birth in ART cycles. They reported that in vitro culture-induced precocious X-chromosome inactivation together with ICSI-induced decrease in the number of trophectoderm cells in female blastocysts may account for the preferential female mortality at early post-implantation stages, resulting in the observed variations in sex ratios at birth in ART cycles [30].

The similar effect was observed in previous study that diverse blastocyst parameter grades were related to the sex ratio of singleton live births in SBT cycles [17]. The proportion of male childbirth from high-quality blastocysts transfer was significantly increased than those from poor-quality blastocysts transfer, which had the trend of elevation with the advance of blastocysts quality. In the present study, we also found that blastocysts with better morphological scores were much more likely to produce male infants, and the trophectoderm grade and ICM grade were both identified as important factors affecting the newborn sex ratio after a single-blastocyst transfer cycle.

The trophectoderm grade A blastocysts had statistically significant association with the sex ratio among newborns conceived via blastocyst transfer,which was supported by M.B. Borgstrøm’s investigation [31]. That is to say, blastocyst morphology was closely related to the sex ratio of livebirth when blastocysts with higher grade trophectoderm were transferred. The contribution of trophectoderm during the implantation stage could be attributed to its impact as a promoter of incubation and endometrial invasion [32]. The study of Ebner found that the morphology of trophectoderm was associated with the sex ratio of offspring, with the rates of implantation, pregnancy, and miscarriage. What matters most was that male blastocysts were 2.53 times more likely than female ones with grade A TE [33]. Previous study have shown that human female embryos consume much higher levels of glucose than male embryos [34], which might reflect the need for more energy in humans to inactivate the second X-chromosome [35] thereby reducing the mitotic progression. Interestingly, we demonstrated that after controlling for the trophoblast grade, ICM grade B was positively correlated with the higher sex ratio among singleton births but not ICM grade A. Thus, disparities in the morphological scores between male and female embryos were likely to reflect some difference in development. Previous studies had different conclusions about the relationship between ICM grade and sex ratio. Lou et al. found that the ICM quality had no obvious effect on the newborn sex ratio after single-blastocyst FET cycles [17]. Kate et al. demonstrated that newly expanded normally fertilized blastocysts had an average of 58.3 ± 8.1 cells on day 5, which increased to 84.4 ± 5.7 and 125.5 ± 19 on days 6 and 7, respectively. The number of TE cells was basically the same on days 5 and 6 (37.9 ± 6.0 and 40.3 ± 5.0, respectively), and doubled on day 7 (80.6 ± 15.2). In contrast, the number of ICM cells doubled between days 5 and 6 (20.4 ± 4.0 and 41.9 ± 5.0, respectively) and remained almost unchanged on day 7 (45.6 ± 10.2) [36]. This was in line with our results, i.e., male embryos grew more rapidly than female ones, contributing to the higher proportion of male blastocysts seen on day 5. However, there were fewer ICM cells than TE cells on day 5, leading to a lower ICM grade. Inversely, the proportion of male embryos declined on day 6 compared to that on day 5; however, the ICM cells increased, which may account for a higher ICM grade. In China, there were studies concerning the ICM grade in relation to the newborn sex ratio, clinical pregnancy rate, live birth rate, and pregnancy loss rate. However, not all results were consistent.In our reproductive center, embryo transfer strategy required that the two best-quality cleavage-stage embryos be chosen for fresh cycle transfer or for cryopreservation on day 3, with the remaining embryos being cultured to the blastocyst stage for possible cryopreservation.

Therefore, we had very few good embryos (grade AA) and a large percentage of “fair” BB and “poor” BC embryos, and the strategy required an ICM grade of grade B or higher for optimal cryosurvival. As we all know, ICM developed into a fetus and TE develops into the placenta eventually,We does not transfer the blastocyst with grade C ICM but TE with grade C can be transferred routinely when the ICM is grade A or B in our center. When an ICM grade was C, the embryos were abandoned. Therefore, we only evaluated the outcomes associated with embryos having an ICM grade of A or B. In this situation, data concerning the ICM had the risk of information bias, which is one of the limitations of the study. However, blastocyst transfer is more advantageous than cleavage-stage embryo transfer in terms of successful implantation rate and pregnancy rates. We prefer high-quality blastocyst transfer to obtain a better clinical pregnancy rate. Although we determined that the transfer of high-quality blastocysts may skew the sex ratio in favor of male embryos, it is difficult to change perceptions regarding popular blastocyst transfer strategies.

In clinical practice, age was a very important factor in considering infertility, which was strongly associated with gametes, embryo, and clinical outcomes. This study was stratified by age of infertile patients, and the results revealed that the sex ratio was significantly affected by blastocyst quality in young patients (≤35 years), which is in agreement with the results of previous studies [1, 3]. Dean et al. referred to female age as being one of the significant factors affecting the probability of having a male birth. They found that a lower age was associated with a higher likelihood of male birth [15]. Bahadur et al. studied a population in the United Kingdom and drew a similar conclusion [37]. This may because of the higher oxidative stress level in advanced maternal age, but male infants had poor survivability when subjected to high oxidative stress injury compared to female infants. Therefore, male infants may be miscarried in early pregnancy among women of older age. Furthermore, our results found that a higher sex ratio was associated with a lower age in men. However, only few studies have reported the effect of paternal age on sex ratio. Jeremy M found that the sex ratio significantly decreased with increasing paternal age and remained so even after adjusting for maternal age in offspring with autism spectrum disorders, suggesting that the reduction in the sex ratio with increasing paternal age consistent with de novo genetic or genomic anomalies, which arise more frequently as older men conceive [37].

This study has some limitations. As the study was only conducted in a single center, the results may not be generalizable. In addition, it is uncertain that the data accurately reflect the real sex ratio at birth finally under the influence of Chinese culture,which is one limitations of this study.The retrospective nature of this study is another limitation. In future, prospective multicenter studies should perform stratified analysis according to maternal or paternal age ≤35 years in order to validate the findings of our study.

## Conclusions

In summary, we demonstrated that the morphological criteria associated with higher-quality blastocysts resulted in a higher sex ratio after SBT. In our study, not only the trophectoderm grade but also the ICM grade was significantly associated with the sex ratio among offspring. The sex ratio increased as the trophectoderm grade increased, whereas a higher ICM grade (grade A) resulted in a lower sex ratio than ICM of grade B, especially in infertile couples in which either partner was below 35 years of age. Our findings may shed light on potential factors underlying the skewed sex ratio among offspring from blastocyst transfer, which provided reference for embryo transfer strategy. In future studies, we would further explore the factors affecting the sex ratio with the aim of maintaining a balanced sex ratio among offspring in IVF/ICSI, without affecting the pregnancy rate.

## Data Availability

within the manuscript and/or Supporting Information files

## List of abbreviations

(IVF): in vitro fertilization
(ICSI-ET): intracytoplasmic sperm injection-embryo transfer
(FET): frozen-thawed embryo transfer
(PSR): primary sex ratio
(SSR): secondary sex ratio
(SBT): single blastocyst transfer
(ICM): inner cell mass
(TE): trophectoderm
(ART): assisted reproductive technologies
(PGT): preimplantation genetic testing
(BMI): body mass index
(DMSO): dimethyl sulfoxide
(OHSS): ovarian hyperstimulation syndrome
(HCG): human chorionic gonadotropin
(HRT): hormone replacement therapy
(SPSS): Statistical Package for Social Sciences

## Declarations

### Ethics approval and consent to participate

The study was carried out after the protocol was approved by the Institutional Review Board of The Third Affiliated Hospital of Zhengzhou University. I confirm that all methods were performed in accordance with the relevant guidelines. The requirement for informed consent was waived by the Institutional Review Board of The Third Affiliated Hospital of Zhengzhou University because of the retrospective nature of the study.

### Consent for publication

Not applicable.

### Availability of data and materials

All data generated or analysed during this study are included in this published article

### Competing interests

The authors declare that they have no competing interests

### Funding

Not applicable.

### Authors’ contributions

Study design: JJH, YCG, and RJY. Data acquisition and analysis: JJH and RJY. Drafting of the manuscript and interpretation: YCG, HL. Revision of the manuscript: LJS, XLW and RJY. The authors read and approved the final manuscript.

## Acknowledgments

The authors would like to thank all of the participants and the staff of the Reproductive Center, the Third Affiliated Hospital of Zhengzhou University, for their selfless assistance.

## Notes

### Competing Interest Statement

The authors have declared no competing interest.

### Author Declarations

The study was carried out after the protocol was approved by the Institutional Review Board of The Third Affiliated Hospital of Zhengzhou University. Informed consent was waived due to retrospective design of the study based on our local ethics committee protocols.

## References

1. Supramaniam PR, Mittal M, Ohuma EO, Lim LN, McVeigh E, Granne I, et al. Secondary sex ratio in assisted reproduction: an analysis of 1 376 454 treatment cycles performed in the UK. Human reproduction open. 2019;2019(4):hoz020. Epub 2019/10/11. doi: 10.1093/hropen/hoz020. PubMed PMID: 31598568.

2. Lin PY, Huang FJ, Kung FT, Wang LJ, Chang SY, Lan KC. Comparison of the offspring sex ratio between cleavage stage embryo transfer and blastocyst transfer. Taiwanese journal of obstetrics & gynecology. 2010;49(1):35–9. Epub 2010/05/15. doi: 10.1016/s1028-4559(10)60006-x. PubMed PMID: 20466290.

3. Hattori H, Kitamura A, Takahashi F, Kobayashi N, Sato A, Miyauchi N, et al. The risk of secondary sex ratio imbalance and increased monozygotic twinning after blastocyst transfer: data from the Japan Environment and Children’s Study. Reproductive biology and endocrinology : RB&E. 2019;17(1):27. Epub 2019/02/24. doi: 10.1186/s12958-019-0471-1. PubMed PMID: 30795788.

4. Gardner DK, Wb S. In vitro culture of human blastocysts. In: Jansen, R.and Mortimer, D., Eds., Towards Reproductive Certainty: Infertility and Genetics Beyond 1999: The Plenary Proceedings of the 11th World Congress on In Vitro Fertilization and Human Reproductive Genetics, Parthenon Press, Pearl River. 1999:378–88.

5. Jacobsen R, Møller H, Mouritsen A. Natural variation in the human sex ratio. Human reproduction (Oxford, England). 1999;14(12):3120–5. Epub 1999/12/22. doi: 10.1093/humrep/14.12.3120. PubMed PMID: 10601107.

6. Hesketh T, Xing ZW. Abnormal sex ratios in human populations: causes and consequences. Proceedings of the National Academy of Sciences of the United States of America. 2006;103(36):13271–5. Epub 2006/08/30. doi: 10.1073/pnas.0602203103. PubMed PMID: 16938885.

7. Wu Z, Viisainen K, Hemminki E. Determinants of high sex ratio among newborns: a cohort study from rural Anhui province, China. Reproductive health matters. 2006;14(27):172–80. Epub 2006/05/23. doi: 10.1016/s0968-8080(06)27222-7. PubMed PMID: 16713892.

8. Fan SL, Xiao CN, Zhang YK, Li YL, Wang XL, Wang L. How does the two-child policy affect the sex ratio at birth in China? A cross-sectional study. BMC Public Health. 2020;20(1):789. doi: 10.1186/s12889-020-08799-y. PubMed PMID: 32460822.

9. De Croo I, Colman R, De Sutter P, Tilleman K. Blastocyst transfer for all? Higher cumulative live birth chance in a blastocyst-stage transfer policy compared to a cleavage-stage transfer policy. Facts, views & vision in ObGyn. 2019;11(2):169–76. Epub 2019/12/12. PubMed PMID: 31824637.

10. Chen H, Lv JQ, Wu XM, Xiao Y, Xi HT, Zhu CF, et al. Blastocyst-stage versus cleavage-stage embryo transfer in the first frozen cycles of OHSS-risk patients who deferred from fresh embryo transfer. Gynecological endocrinology : the official journal of the International Society of Gynecological Endocrinology. 2015;31(9):698–701. Epub 2015/07/21. doi: 10.3109/09513590.2015.1062858. PubMed PMID: 26190533.

11. Zhu L, Xi Q, Zhang H, Li Y, Ai J, Jin L. Blastocyst culture and cryopreservation to optimize clinical outcomes of warming cycles. Reproductive biomedicine online. 2013;27(2):154–60. Epub 2013/06/19. doi: 10.1016/j.rbmo.2013.04.006. PubMed PMID: 23769665.

12. Hur YS, Ryu EK, Song SH, Yoon SH, Lim KS, Lee WD, et al. A retrospective study of single frozen-thawed blastocyst transfer. Clinical and experimental reproductive medicine. 2016;43(2):106–11. Epub 2016/07/01. doi: 10.5653/cerm.2016.43.2.106. PubMed PMID: 27358829.

13. Maheshwari A, Hamilton M, Bhattacharya S. Should we be promoting embryo transfer at blastocyst stage? Reproductive biomedicine online. 2016;32(2):142–6. Epub 2015/12/18. doi: 10.1016/j.rbmo.2015.09.016. PubMed PMID: 26673100.

14. Monteleone PA, Petersen PG, Peregrino PF, Miorin J, Gomes AP, Fujii MG, et al. Should single embryo transfer be used in patients with any kind of infertility factor? Preliminary outcomes. JBRA assisted reproduction. 2019;23(3):200–4. Epub 2019/03/27. doi: 10.5935/1518-0557.20190006. PubMed PMID: 30912516.

15. Dean JH, Chapman MG, Sullivan EA. The effect on human sex ratio at birth by assisted reproductive technology (ART) procedures--an assessment of babies born following single embryo transfers, Australia and New Zealand, 2002-2006. BJOG : an international journal of obstetrics and gynaecology. 2010;117(13):1628–34. Epub 2010/09/30. doi: 10.1111/j.1471-0528.2010.02731.x. PubMed PMID: 20875033.

16. Sharara FI, Abdo G. Incidence of monozygotic twins in blastocyst and cleavage stage assisted reproductive technology cycles. Fertility and sterility. 2010;93(2):642–5. Epub 2009/02/17. doi: 10.1016/j.fertnstert.2008.12.130. PubMed PMID: 19217097.

17. Lou H, Li N, Zhang X, Sun L, Wang X, Hao D, et al. Does the sex ratio of singleton births after frozen single blastocyst transfer differ in relation to blastocyst development? Reproductive biology and endocrinology : RB&E. 2020;18(1):72. doi: 10.1186/s12958-020-00623-x. PubMed PMID: 32669110.

18. Chen X, Zhang J, Wu X, Cao S, Zhou L, Wang Y, et al. Trophectoderm morphology predicts outcomes of pregnancy in vitrified-warmed single-blastocyst transfer cycle in a Chinese population. Journal of assisted reproduction and genetics. 2014;31(11):1475–81. Epub 2014/08/16. doi: 10.1007/s10815-014-0317-x. PubMed PMID: 25123128.

19. Sun LJ, Zhao B, Hu JJ. Application of three kinds of ovulation induction in in vitro fertilization/intracytoplasmic sperm injection patients with poor ovarian response. Chin J Reprod Contracep. 2018;38(03):186–91.

20. Wang Ll, Sun LJ, Feng YY. A comparative study of ovulation promotion effect between three kinds of down regulation long protocol in infertile IVF/ICSI patients with normal ovarian function. Prog Obstet Gynecol. 2019;28(08):597–601.

21. Papanikolaou EG, D’Haeseleer E, Verheyen G, Van de Velde H, Camus M, Van Steirteghem A, et al. Live birth rate is significantly higher after blastocyst transfer than after cleavage-stage embryo transfer when at least four embryos are available on day 3 of embryo culture. A randomized prospective study. Human reproduction (Oxford, England). 2005;20(11):3198–203. Epub 2005/08/02. doi: 10.1093/humrep/dei217. PubMed PMID: 16055454.

22. Schoolcraft WB, Gardner DK, Lane M, Schlenker T, Hamilton F, Meldrum DR. Blastocyst culture and transfer: analysis of results and parameters affecting outcome in two in vitro fertilization programs. Fertility and sterility. 1999;72(4):604–9. Epub 1999/10/16. doi: 10.1016/s0015-0282(99)00311-8. PubMed PMID: 10521095.

23. Herbemont C, Chekroune S, Bonan S, Cedrin-Durnerin I, Vivot A, Sonigo C, et al. Impact of post-warming culture duration on clinical outcomes of vitrified good-quality blastocyst transfers: a prospective randomized study. Fertility and sterility. 2018;110(7):1290–7. Epub 2018/12/07. doi: 10.1016/j.fertnstert.2018.07.1153. PubMed PMID: 30503128.

24. Bu Z, Chen ZJ, Huang G, Zhang H, Wu Q, Ma Y, et al. Live birth sex ratio after in vitro fertilization and embryo transfer in China--an analysis of 121,247 babies from 18 centers. PloS one. 2014;9(11):e113522. Epub 2014/11/21. doi: 10.1371/journal.pone.0113522. PubMed PMID: 25412419.

25. Fernando D, Halliday JL, Breheny S, Healy DL. Outcomes of singleton births after blastocyst versus nonblastocyst transfer in assisted reproductive technology. Fertility and sterility. 2012;97(3):579–84. Epub 2012/01/28. doi: 10.1016/j.fertnstert.2011.12.032. PubMed PMID: 22281036.

26. Lin PY, Huang FJ, Kung FT, Wang LJ, Chang SY, Lan KC. Comparison of the offspring sex ratio between fresh and vitrification-thawed blastocyst transfer. Fertility and sterility. 2009;92(5):1764–6. Epub 2009/06/16. doi: 10.1016/j.fertnstert.2009.05.011. PubMed PMID: 19523616.

27. Wikland M, Hardarson T, Hillensjö T, Westin C, Westlander G, Wood M, et al. Obstetric outcomes after transfer of vitrified blastocysts. Human reproduction (Oxford, England). 2010;25(7):1699–707. Epub 2010/05/18. doi: 10.1093/humrep/deq117. PubMed PMID: 20472913.

28. Wang M, Liu X, Zhang H, Li L, Liu R, Zhang H, et al. Associated factors of secondary sex ratio of offspring in assisted reproductive technology: a cross-sectional study in Jilin Province, China. BMC pregnancy and childbirth. 2020;20(1):666. Epub 2020/11/06. doi: 10.1186/s12884-020-03373-1. PubMed PMID: 33148206.

29. Luke B, Brown MB, Grainger DA, Baker VL, Ginsburg E, Stern JE. The sex ratio of singleton offspring in assisted-conception pregnancies. Fertility and sterility. 2009;92(5):1579–85. Epub 2008/10/28. doi: 10.1016/j.fertnstert.2008.08.107. PubMed PMID: 18950756.

30. Tarín JJ, García-Pérez MA, Hermenegildo C, Cano A. Changes in sex ratio from fertilization to birth in assisted-reproductive-treatment cycles. Reproductive biology and endocrinology : RB&E. 2014;12:56. Epub 2014/06/25. doi: 10.1186/1477-7827-12-56. PubMed PMID: 24957129.

31. Borgstrøm MB, Kesmodel US, Klausen TW, Danielsen AK, Thomsen T, Gabrielsen A, et al. Developmental stage and morphology of the competent blastocyst are associated with sex of the child but not with other obstetric outcomes: a multicenter cohort study. Human reproduction (Oxford, England). 2021;37(1):119–28. Epub 2022/01/06. doi: 10.1093/humrep/deab242. PubMed PMID: 34986219.

32. Ahlström A, Westin C, Reismer E, Wikland M, Hardarson T. Trophectoderm morphology: an important parameter for predicting live birth after single blastocyst transfer. Human reproduction (Oxford, England). 2011;26(12):3289–96. Epub 2011/10/06. doi: 10.1093/humrep/der325. PubMed PMID: 21972253.

33. Ebner T, Tritscher K, Mayer RB, Oppelt P, Duba HC, Maurer M, et al. Quantitative and qualitative trophectoderm grading allows for prediction of live birth and gender. Journal of assisted reproduction and genetics. 2016;33(1):49–57. Epub 2015/11/18. doi: 10.1007/s10815-015-0609-9. PubMed PMID: 26572782.

34. Gardner DK, Wale PL. Analysis of metabolism to select viable human embryos for transfer. Fertility and sterility. 2013;99(4):1062–72. Epub 2013/01/15. doi: 10.1016/j.fertnstert.2012.12.004. PubMed PMID: 23312219.

35. Epstein CJ, Smith S, Travis B, Tucker G. Both X chromosomes function before visible X-chromosome inactivation in female mouse embryos. Nature. 1978;274(5670):500–3. Epub 1978/08/03. doi: 10.1038/274500a0. PubMed PMID: 672979.

36. Hardy K, Handyside AH, Winston RM. The human blastocyst: cell number, death and allocation during late preimplantation development in vitro. Development (Cambridge, England). 1989;107(3):597–604. Epub 1989/11/01. PubMed PMID: 2612378.

37. Anello A, Reichenberg A, Luo X, Schmeidler J, Hollander E, Smith CJ, et al. Brief report: parental age and the sex ratio in autism. Journal of autism and developmental disorders. 2009;39(10):1487–92. Epub 2009/05/20. doi: 10.1007/s10803-009-0755-y. PubMed PMID: 19452267.

